# Screening for Osteoporosis: A Draft Update of Systematic Review Adapted from the USPSTF Review for the Japan Preventive Services Task Force

**DOI:** 10.1101/2025.06.26.25330314

**Authors:** Takao kaneko, Yuichi Isaji, Hiroki Sato, Satoshi Miike, Hironori Yamada, Ryohei Oda, Toshiya Sakoda

## Abstract

**Background:** Fragility fractures are a major public health concern among older adults in Japan. In 2024, the U.S. Preventive Services Task Force (USPSTF) released a draft update of its evidence review on screening for osteoporosis. We conducted a systematic review, commissioned by the Japan Preventive Services Task Force (JPPSTF), to update and apply the USPSTF draft by incorporating Japanese-language literature and additional recent international evidence.

**Methods:** This review followed the USPSTF analytic framework. We evaluated evidence on the effectiveness of screening; the predictive and diagnostic accuracy of clinical risk assessment tools; the predictive accuracy of bone mineral density (BMD) testing; the potential harms of screening; and the benefits and harms of pharmacologic treatments approved in Japan. Literature searches were conducted by the International Medical Information Center (IMIC). PubMed and the Cochrane Library were searched for studies published from November 11, 2022, to November 26 2024, and Ichushi-Web (Japan Medical Abstracts Society) was searched without publication date restriction. Searches were limited to English and Japanese. Study selection, data extraction, and risk of bias assessment were conducted independently by two or more reviewers for each key question, and disagreements were resolved through discussion.Findings from newly identified studies were qualitatively synthesized alongside the USPSTF review.

**Results:** Osteoporosis screening was generally associated with a modest reduction in hip and major osteoporotic fractures among postmenopausal women, particularly those aged 65 years or older. Risk assessment tools showed moderate predictive accuracy; diagnostic accuracy varied. Harms of screening were infrequently reported. Pharmacologic treatments approved in Japan were associated with fracture risk reduction, with no consistent evidence of serious harms. These findings were broadly consistent with those of the USPSTF review.

**Conclusions:** This systematic review provides an updated and objective evaluation of the internal validity of current evidence on osteoporosis screening and treatment in primary care settings in Japan.

## Background

Fragility fractures are a major and growing public health concern among older adults in Japan^1–3^. The primary purpose of osteoporosis screening is to identify individuals who may benefit from pharmacologic treatment to reduce the incidence and morbidity of such fractures.While internationally referenced, evidence-based recommendations, such as those from the U.S. Preventive Services Task Force (USPSTF), have guided global practice on osteoporosis screening, previous reviews^4^ have primarily relied on English-language literature and have included only limited evidence from studies conducted in Japan. In 2024, the USPSTF issued a draft^5^ update of its evidence review, reaffirming its recommendation to conduct screening in women aged 65 years and older, and in younger postmenopausal women who are at increased risk. However, the evidence remained insufficient to assess the balance of benefits and harms of screening in men.

To address this gap, the Japan Preventive Services Task Force (JPPSTF) commissioned the Literature Review Team for Osteoporosis Screening to conduct a systematic review that incorporates Japanese-language literature in addition to newly published English-language studies. The team evaluated the internal validity of evidence on the effectiveness, accuracy, and potential harms of osteoporosis screening and pharmacologic interventions, with particular attention to studies relevant to the Japanese population.

## Materials and Methods

### Scope of the Review

This review followed the PRISMA 2020 (Preferred Reporting Items for Systematic Reviews and Meta-Analyses) statement^6^ for reporting systematic reviews. The review was guided by the analytic framework shown in Figure 1. The framework was adapted from the 2024 draft evidence review by the U.S. Preventive Services Task Force (USPSTF) and revised to reflect the Japanese context.

**Figure 1.**
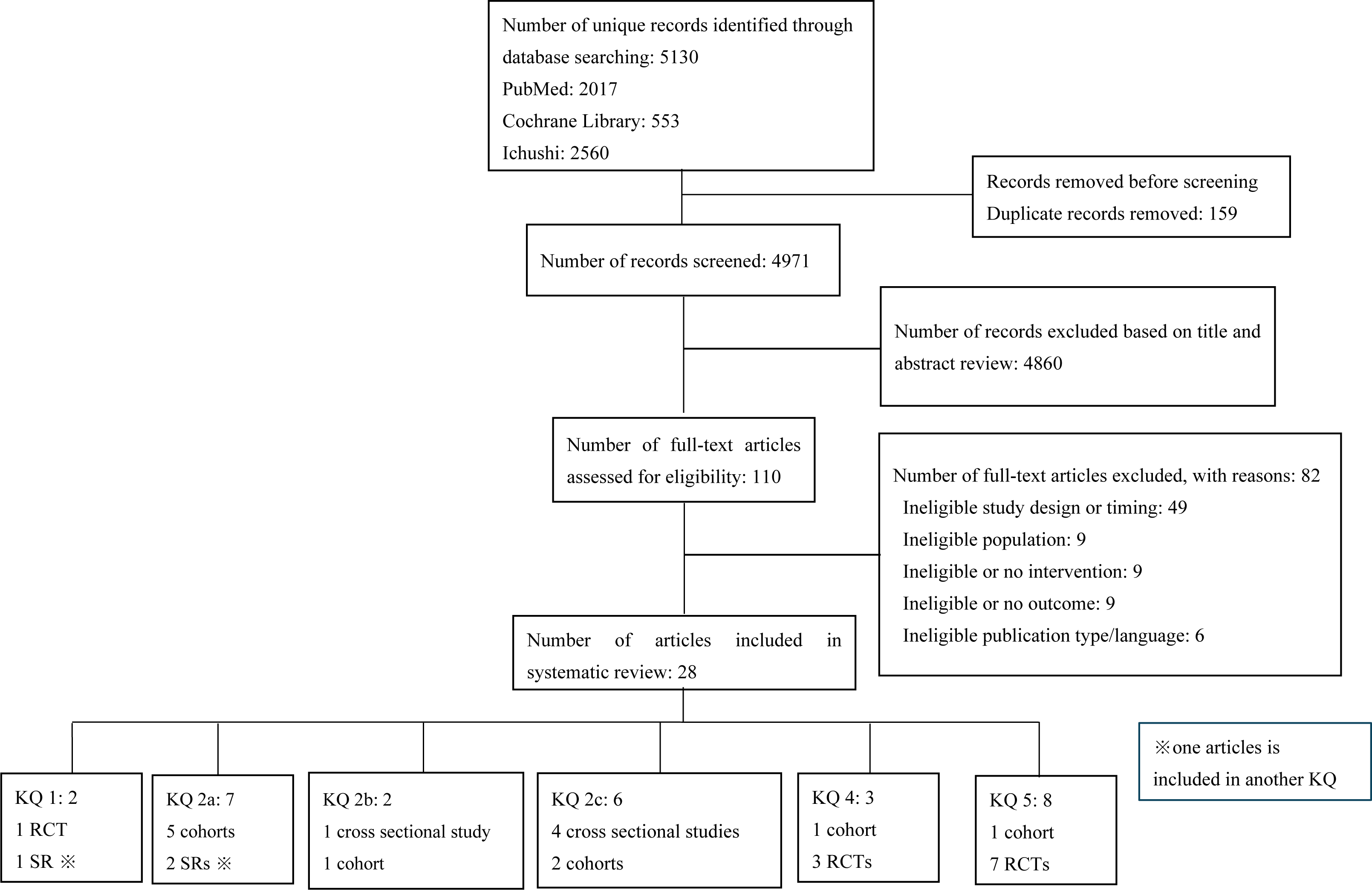
Literature Flow Diagram.

### Data Sources and Searches

Literature searches were conducted by the International Medical Information Center (IMIC) on November 26, 2024. PubMed and the Cochrane Library were searched for studies published in English or Japanese between November 11, 2022, and November 26, 2024. Ichushi-Web (Japan Medical Abstracts Society) was searched without restriction on publication year. Details of the search strategies are provided in the Supplement.

### Study Selection

Titles and abstracts were screened using Rayyan (https://www.rayyan.ai), followed by full-text review, by two or more independent reviewers using prespecified eligibility criteria. Disagreements were resolved through discussion. Reviewer assignments were structured by the Literature Review Team for Osteoporosis Screening to maintain methodological independence across key questions. Eligibility criteria were defined separately for each key question (KQ). Depending on the question, eligible study designs included randomized controlled trials, controlled cohort studies, diagnostic accuracy studies, and systematic reviews. For KQ1, studies evaluating the effectiveness of osteoporosis screening compared with no screening in adults without known osteoporosis or prior fragility fracture were included. For KQs 2a–2d, studies assessing the predictive or diagnostic performance of risk assessment tools (e.g., FRAX, OSTA) or BMD testing were included if they were externally validated in at least one independent cohort. For KQ3, studies examining harms such as overdiagnosis, labeling, or unnecessary treatment were eligible. For KQs 4 and 5, studies evaluating pharmacologic agents approved for osteoporosis treatment in Japan were eligible. Studies focused on secondary osteoporosis, prior fragility fractures, or treatment-refractory populations were excluded.

### Data Extraction and Quality Assessment

Relevant data (e.g., study design, population, intervention, comparator, outcomes, and setting) were extracted independently by two or more reviewers using a standardized form. Discrepancies were resolved through discussion.

Risk of bias was assessed using appropriate tools: RoB 2^7^ for randomized controlled trials, ROBINS-I^8^ for observational studies, QUADAS-2^9^ for diagnostic accuracy studies, PROBAST^10^ for prediction model studies, and ROBIS^10^ for systematic reviews. Each study was independently assessed by two or more reviewers. Any differences in judgment were resolved by consensus.

### Data Synthesis and Analysis

Findings from newly identified studies were qualitatively synthesized alongside the USPSTF review. For each key question, results were summarized in descriptive and tabular formats, with attention to the direction and consistency of findings, study design, and methodological quality.

A meta-analysis was not performed, as a relatively recent draft^5^ systematic review by the USPSTF was already available. In addition, this decision reflected the need to complete the current review within a limited timeframe.

## Results

A total of 28 studies met eligibility criteria and were included in this systematic review (Figure1). One study was relevant to two key questions, and thus was counted under both questions in the presentation of results. The distribution of studies by key question was as follows: 2 studies for KQ1, 15 studies for KQ2 (subdivided into KQ2a: 7 studies, KQ2b: 2 studies, KQ2c: 6 studies), none for KQ3, 4 studies for KQ4, and 8 studies for KQ5.The results of the included studies were synthesized and presented according to each KQ, as outlined below.

KQ1: Effectiveness of Osteoporosis Screening (Table 1)

**Table 1.**
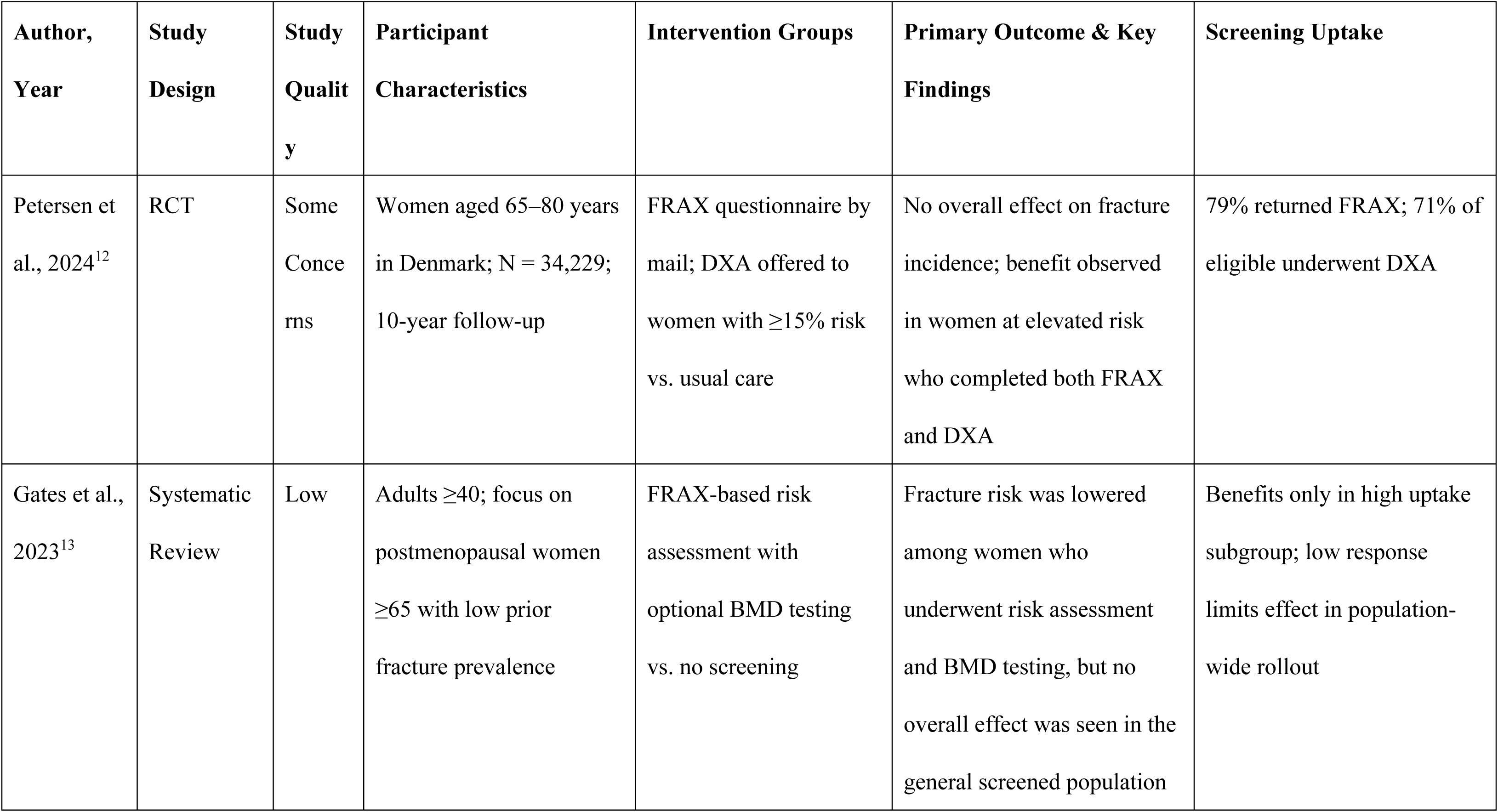
Characteristics of Included Studies for KQ1.

| **Author, Year** | **Study Design** | **Study Quality** | **Participant Characteristics** | **Intervention Groups** | **Primary Outcome & Key Findings** | **Screening Uptake** |
| --- | --- | --- | --- | --- | --- | --- |
| Petersen et al., 2024<sup>12</sup> | RCT | Some Concerns | Women aged 65–80 years in Denmark; N = 34,229; 10-year follow-up | FRAX questionnaire by mail; DXA offered to women with $\geq 15\%$ risk vs. usual care | No overall effect on fracture incidence; benefit observed in women at elevated risk who completed both FRAX and DXA | 79% returned FRAX; 71% of eligible underwent DXA |
| Gates et al., 2023<sup>13</sup> | Systematic Review | Low | Adults $\geq 40$; focus on postmenopausal women $\geq 65$ with low prior fracture prevalence | FRAX-based risk assessment with optional BMD testing vs. no screening | Fracture risk was lowered among women who underwent risk assessment and BMD testing, but no overall effect was seen in the general screened population | Benefits only in high uptake subgroup; low response limits effect in population-wide rollout |

Two studies^12,13^ met the inclusion criteria for evaluating the effectiveness of osteoporosis screening in reducing the incidence of fractures or improving clinical outcomes in adults without prior diagnosis of osteoporosis or fragility fractures.

The first was the ROSE randomized trial, which implemented a two-step screening strategy involving a mailed, self-administered FRAX questionnaire to identify women at elevated fracture risk (FRAX ≥15%), followed by DXA scanning for those meeting the risk threshold. A reduction in fracture incidence was observed exclusively among women who both completed the questionnaire and underwent DXA, indicating that the effectiveness of screening was confined to individuals at moderate to high risk who fully participated in the screening pathway.

The second study was a systematic review conducted for the Canadian Task Force, which followed by BMD testing in high-risk individuals. The review found moderate-certainty evidence that reduce the risk of hip and clinical fragility fractures among women. In contrast, screening offered to the general population without sufficient participation did not yield benefit. The review also noted the potential for overdiagnosis in individuals classified as high risk by FRAX, despite having a low actual likelihood of sustaining fractures.

KQ2: Accuracy of Screening Strategies in Identifying High-Risk Individuals

KQ2a: Performance of Clinical Risk Assessment Tools (FRAX, OSTA) for Predicting Hip and Major Osteoporotic Fractures (Table 2)

**Table 2.** Characteristics of Included Studies for KQ2a.

| **Author,**<br>**Year; Study**<br>**Quality** | **Cohort**<br>**Name;**<br>**Country** | **Cohort Description** | **N; %**<br>**Female** | **Mean Age (SD)** | **Additional information** |
| --- | --- | --- | --- | --- | --- |
| Crandall et al., 2023<sup>14</sup>;<br>Moderate | WHI;<br>United States | Postmenopausal women aged 50–64, stratified by race/ethnicity | 67,169;<br>100% | 57.8 (4.1) | Evaluated FRAX (without BMD) and OST stratified by race/ethnicity. For prediction of 10-year major osteoporotic fracture (MOF), FRAX AUCs: Asian 0.65, Black 0.55, Hispanic 0.61, White 0.59. FRAX underestimated MOF risk in Black women and overestimated it in Asian women. |
| Fink et al., 2023<sup>15</sup>;<br>Some Concerns | WHI subset;<br>United States | Postmenopausal women aged 50–79 stratified by race/ethnicity | 84,455;<br>100% | Mean age 63.2 (7.3), varied by race/ethnicity (e.g., Asian 61.3, White 63.9) | Compared FRAX and Garvan models by race/ethnicity; outcomes: MOF and hip fracture; AUCs ranged 0.58–0.76 |
| Freitas et al.,<br>2024<sup>16</sup>;<br>Moderate | SPAH;<br>Brazil | Community-dwelling<br>adults aged $\geq 65$ years | 705;<br>63% | 72.6 (4.6) | Assessed FRAX and NOGG (±BMD); AUCs: NVF<br>0.76, MOF 0.67; FRAX without BMD performed<br>comparably or better |
| Gates et al.,<br>2023<sup>13</sup>; Low | Multiple<br>Canadian<br>cohorts;<br>Canada | Adults aged $\geq 40$ years<br>from population-based<br>cohorts including<br>CaMos, evaluated for<br>calibration of FRAX<br>with and without<br>BMD | 67,611;<br>majority<br>female | Median age 67 (IQR 57–75) | Calibration of FRAX-Canada with and without BMD |
| Leslie et al.,<br>2023<sup>17</sup>;<br>Moderate | Manitoba<br>BMD<br>Registry;<br>Canada | Postmenopausal<br>women referred for<br>BMD testing; used<br>registry-linked<br>fracture outcomes | 45,185;<br>100% | Age-stratified (e.g., $<65$, $65-74$, $\geq 75$ years) | Evaluated FRAX adjustment using renormalized L1-only TBS; assessed improvement in fracture risk<br>prediction and calibration |
| Pluskiewicz et al., 2020<sup>18</sup>; Moderate | GO Study; Poland | Postmenopausal women attending osteoporosis clinic; no prior fracture treatment | 457; 100% | 64.2 (5.9) | Compared predictive performance of FRAX, Garvan, and POL-RISK for MOF and any fracture; AUCs ranged from 0.63 to 0.72 |
| Wu et al., 2023<sup>19</sup>; Moderate | UK Biobank; United Kingdom | Postmenopausal women of European ancestry | 25,772; 100% | 63.2 (7.3) | Used Bayesian-optimized machine learning models incorporating genetic risk scores to improve osteoporotic fracture prediction |

Seven studies^13–19^ evaluated tools such as FRAX and OSTA. Overall, these tools demonstrated moderate to good discriminative ability for predicting major osteoporotic fractures (MOF), especially when BMD was incorporated into the FRAX algorithm. Reported area under the receiver operating characteristic curve (AUC) values generally ranged from 0.65 to 0.78, suggesting fair predictive capacity. In many of the studies that conducted external validation, FRAX with BMD was shown to have higher predictive accuracy than FRAX without BMD or simpler tools like OSTA. Notably, several studies emphasized variation in performance across age strata, with diminished discrimination in very elderly populations, likely due to competing risks. Study settings also varied, encompassing both community-dwelling older adults and health checkup cohorts. One study explored machine learning-enhanced FRAX models, but generalizability was limited. Calibration metrics were infrequently reported, and thresholds for intervention varied.

KQ2b: Accuracy of Bone Mass Measurement Tools in Predicting Hip and Major Osteoporotic Fractures (Table 3)

**Table 3.** Characteristics of Included Studies for Predictive Accuracy of Bone Mineral Density (BMD) Alone for Fracture (Key Question 2b)

| **Author, Year**<br>**Country**<br>**Study**<br>**Quality** | **Population (setting &**<br>**key eligibility)** | **Mean Age**<br>**± SD** | **N (% Female)** | **Mean BMD ± SD (g cm<sup>-2</sup>) &**<br>**Prevalence of Osteoporosis** | **Reference-Test Details (DXA)** |
| --- | --- | --- | --- | --- | --- |
| Lee et al.,<br>2024<sup>20</sup><br>Korea<br>Moderate | Rural adults ≥50 y in<br>Chungju Metabolic<br>Disease Cohort;<br>community health-exam<br>program; no exclusion<br>for comorbidity except<br>missing data | 64.3 ± 9.1 | 5,126 (61.9 %) | Lumbar spine BMD 0.90 ± 0.20;<br>femoral-neck −0.7 ± 1.2 T-score;<br>osteoporosis 32.3 % overall (men 7.6 %, women 41.3 %) | Hologic QDR-4500C DXA;<br>lumbar spine (L1-L4), femoral<br>neck & total hip; CV 1.23 %<br>(spine), 2.3 % (neck), 1.0 % (hip) |
| Zhang et al.,<br>2024<sup>21</sup><br>China<br>Moderate | Women ≥50 y who had<br>lumbar X-ray + DXA<br>within 3 mo at 3 tertiary<br>centres; retrospective | 65.7 ± 9.0 | 1,325 (100 %) | Training: mean BMD 0.90 ± 0.20; T-score −1.81 ± 1.57; osteoporosis 35.9 %<br>Test: mean BMD 0.88 ± 0.18; T-score −1.90 ± 1.53; osteoporosis 37.9 % | GE Lunar Prodigy DXA; antero-posterior lumbar spine (L1-L4);<br>WHO criteria (T-score ≤ −2.5)<br>used as reference standard |

Two studies^20,21^ evaluated the predictive performance of different BMD assessment modalities in identifying individuals at elevated risk for hip or major osteoporotic fractures. Together, these studies suggest that BMD measurements—whether obtained through conventional DXA or emerging radiographic approaches—may be useful for predicting osteoporotic fractures. However, model performance may vary by population, modality, and anatomical site assessed.

KQ2c: Diagnostic Accuracy of Clinical Risk Assessment Tools for Osteoporosis (Table 4)

**Table 4.** Characteristics of Included Studies for Diagnostic Accuracy of Risk Assessment Instruments (Key Question 2c)

| **Author,**<br>**Year**<br>**Country**<br>**ROB** | **Population** | **Mean Age**<br>**(SD)** | **N (%) Female** | **Prevalence of Osteoporosis** | **Reference Test Details** |
| --- | --- | --- | --- | --- | --- |
| Cheung et al., 2023<sup>22</sup><br>Hong Kong<br>Moderate | Postmenopausal women and men $\geq 50$ years from the Hong Kong Osteoporosis Study; community-dwelling | Development / Validation cohorts;<br>Female: 62.8 (6.1) / 63.1 (6.0)<br>Male: 70.3 (4.2) / 71.1 (4.3) | 4747 (data split into validation and development cohorts, % female not specified) | Women <65: not reported; Women $\geq 65$: not reported; Men: not reported<br>PPV for osteoporosis: Women <65: 40.6%, Women $\geq 65$: 59.4%, Men: 19.4% | DXA at femoral neck; T-score $\leq -2.5$; Hologic QDR 4500 plus used |
| Hsieh et al.,<br>2022<sup>23</sup><br>Taiwan<br>Moderate | Adults $\geq 50$ years in<br>rural communities in<br>Taiwan; community-dwelling, cross-<br>sectional | $75.0 \pm 7.1$ | 624 (77.5%) | Men: 12.2%, Women: 51.8% | DXA (Hologic Discovery Wi);<br>lumbar spine and femoral neck T-score $\leq -2.5$ |
| Leeyaphan et<br>al., 2021<sup>24</sup><br>Thailand<br>Moderate | Outpatients with heart<br>failure aged $\geq 60$ years<br>from Ramathibodi<br>Hospital | $72.5 \pm 7.5$ | 185 (56.9%) | 43.6% | DXA (GE Lunar Prodigy); lumbar<br>spine and femoral neck |
| Numazawa et<br>al., 2022<sup>25</sup><br>Multi-<br>country<br>(Asia)<br>Moderate | Community-dwelling<br>Asians $\geq 65$ years from<br>5 Asian<br>countries/regions in<br>the Asian Working<br>Group for Sarcopenia<br>study | $75.1 \pm 6.1$ | 1,321 (55.9%) | Not directly reported; validation of risk<br>model predicting osteoporosis | DXA; details of anatomical site<br>and device not reported |
| Tan et al.,<br>2022<sup>26</sup><br>China<br>Moderate | Hospital-based elderly<br>patients (≥65 y) with<br>type 2 diabetes<br>mellitus | 70.3 ± 6.8 | 229 (58.6%) | 35.1% | DXA (lumbar spine and femoral<br>neck); manufacturer not stated |
| Yamamoto et<br>al., 2013<sup>27</sup><br>Japan<br>Moderate | General women<br>undergoing municipal<br>osteoporosis screening<br>(ages 40–70);<br>Kanazawa City | Not directly<br>reported;<br>stratified by<br>5-year<br>groups | 3,071 (100%) | 36.8% had YAM <80% | DXA or MD method; YAM<br><80% used as proxy for<br>osteoporosis |

Six studies^22–27^ evaluated the diagnostic accuracy of clinical risk assessment tools, either alone or in combination with BMD testing, for identifying individuals with osteoporosis. Across studies, combining clinical risk scores (such as FRAX, OSTA, or locally developed models) with BMD significantly improved diagnostic performance compared to risk scores alone. This integration was especially effective for identifying individuals with intermediate risk profiles, where clinical information alone yielded ambiguous results.

Several studies emphasized that models calibrated to specific populations (e.g., Japanese or East Asian cohorts) demonstrated improved discrimination over internationally derived tools. Additionally, combining multiple clinical factors—such as age, BMI, diabetes, or cardiovascular disease history— with BMD enhanced sensitivity and specificity. However, variability in T-score reference standards and site-specific thresholds (e.g., femoral neck vs. lumbar spine) limited cross-study comparability. Importantly, these findings suggest the usefulness of integrated approaches in clinical practice, particularly in settings where full access to DXA is limited.Incorporating tailored risk models with selectively performed imaging or simplified screening tools may improve the efficiency of case-finding in community-based screening programs.

KQ4: Effectiveness of Pharmacologic Interventions (Table 5)

**Table 5.**
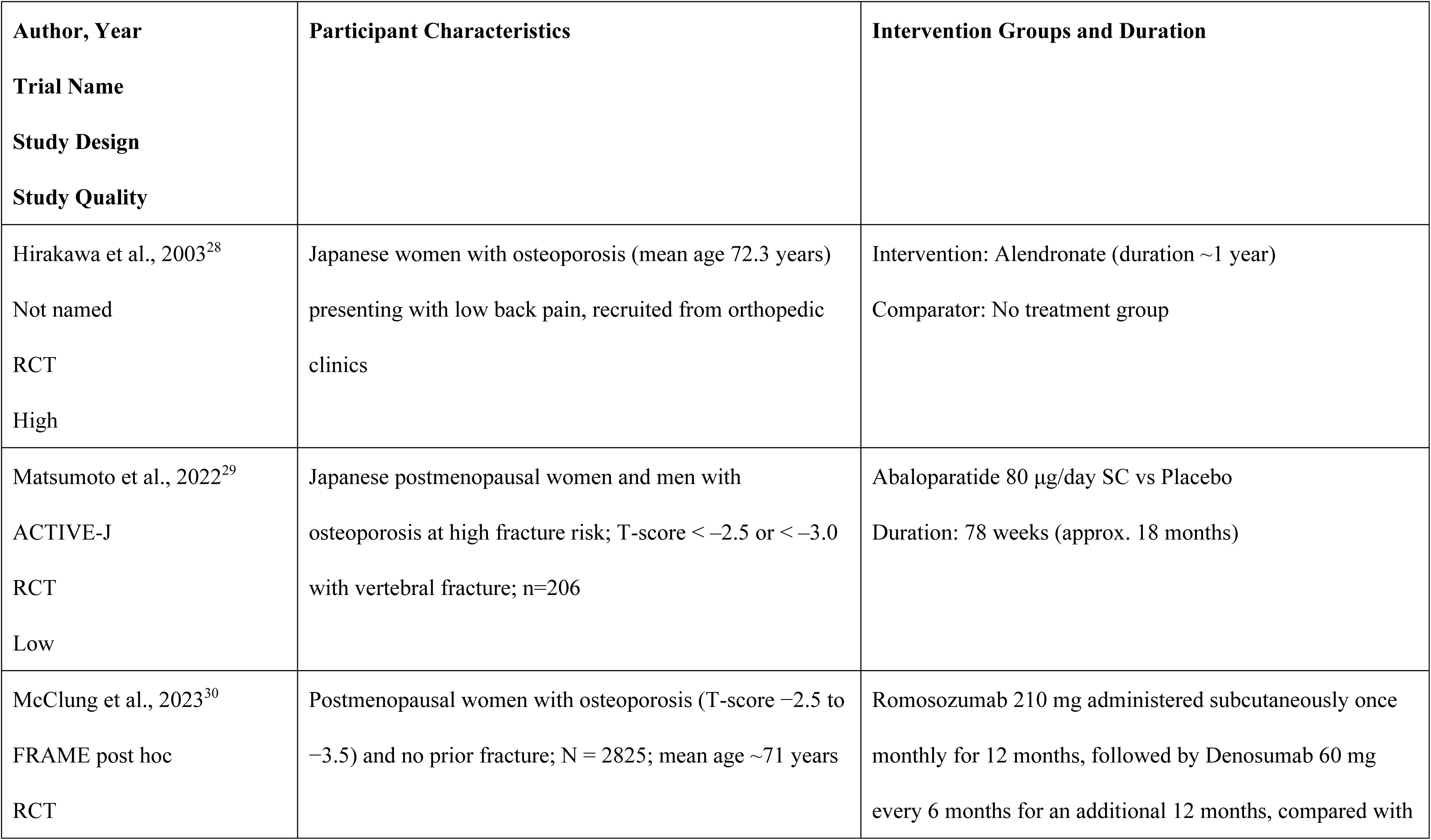
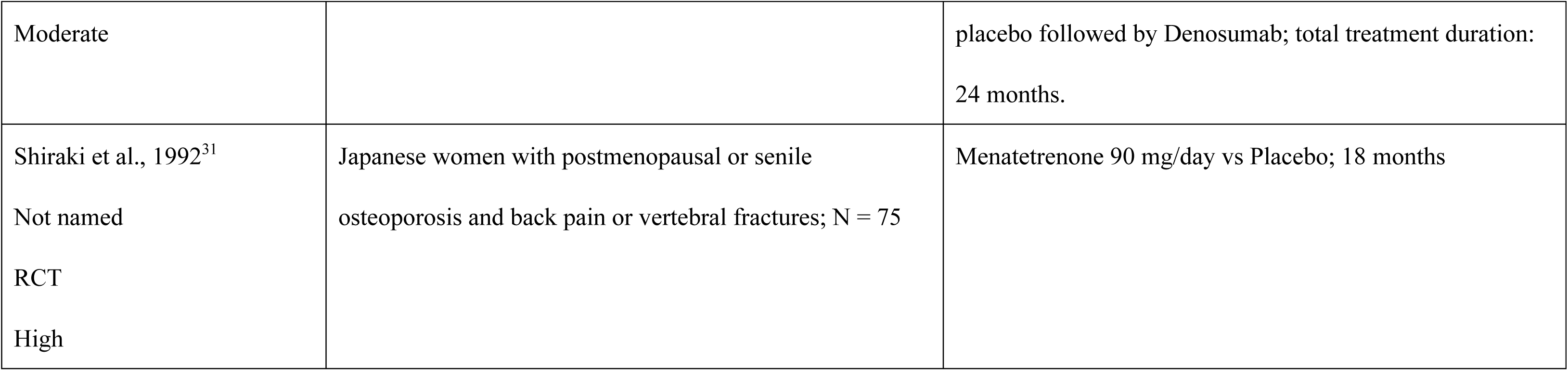
Characteristics of Included Randomized, Controlled Trials for of Treatment (Key Questions 4)

Four randomized controlled trials (RCTs) ^28–31^ evaluated pharmacologic treatments for osteoporosis, including alendronate, menatetrenone, abaloparatide, and sequential romosozumab followed by denosumab. One trial of alendronate compared with no treatment showed a lower vertebral fracture rate in the treated group over one year, though risk of bias was critical. A study of menatetrenone also reported reduced fracture rates, but statistical significance was unclear, and the study had high risk of bias. In contrast, trials of abaloparatide and romosozumab demonstrated statistically significant reductions in vertebral and nonvertebral fractures, with accompanying improvements in bone mineral density. These two trials were judged to have low or moderate risk of bias, respectively.

Overall, recent large-scale trials of anabolic and antiresorptive agents provided more robust evidence, whereas earlier studies suggested benefit but were methodologically limited.Four RCTs^27–30^ conducted evaluated the efficacy of pharmacologic agents approved for osteoporosis management, including bisphosphonates (alendronate), selective estrogen receptor modulators (SERMs), parathyroid hormone analogs (abaloparatide), and RANK ligand inhibitors (denosumab, romosozumab). All trials demonstrated statistically significant reductions in vertebral fracture incidence compared with placebo or standard care. One trial of abaloparatide also showed significant improvement in lumbar spine and hip BMD over 18 months. Romosozumab, followed by denosumab, was associated with sustained antifracture efficacy and greater BMD gains than sequential denosumab alone. Notably, the degree of BMD increase in the romosozumab-to-denosumab sequence was more than twice that of placebo-to-denosumab. These findings suggest that early intervention with osteoanabolic agents may be beneficial for improving bone mineral density and preventing fractures in patients at very high risk of fracture.The overall risk of bias was low in three trials and moderate in one due to incomplete outcome data.

KQ5: Harms of Pharmacologic Interventions (Table 6)

**Table 6.** Characteristics of Included Randomized, Controlled Trials for Harms of Treatment (Key Questions 5)

| **Drug** | **Denosumab** | **Abaloparatide** | **Teriparatide** | **Zoledronic acid** | **Romosozumab** |
| --- | --- | --- | --- | --- | --- |
| Study | Gu et al 2023<sup>32</sup>,<br>Jiang et al 2024<sup>33</sup>,<br>Zhang et al 2023<sup>34</sup> | Czewinski et al 2022<sup>35</sup>,<br>Mastumoto et al 2023<sup>36</sup> | Kontogeorgos et al 2022<sup>37</sup> | Rubin et al 2020<sup>38</sup> | Lane et al 2024<sup>39</sup> |
| Adverse<br>Events | 1. Infections<br>Upper respiratory<br>infection, Urinary tract<br>infection, White blood<br>cells urine positive,<br>Zoster, Pharyngitis,<br>Periodontitis, Periodontal<br>disease<br>2. Musculoskeletal<br>Symptoms<br>Arthralgia, Pain in | 1. Infections and<br>Inflammatory<br>Conditions<br>Nasopharyngitis, Upper<br>respiratory tract<br>inflammation,<br>Bronchitis,<br>Conjunctivitis, Cystitis<br>2. Musculoskeletal<br>Arthralgia, Pain in<br>extremity, Back Pain | 1. Musculoskeletal<br>pain in the limbs<br>2. Neurological and<br>General<br>dizziness, fatigue<br>3. Cardiovascular<br>myocardial infarction | 1. Cardiovascular<br>Atrial fibrillation,<br>Arrhythmias, Myocardial<br>infarction, MI seq, Heart<br>failure, Cardiovascular<br>mortality<br>2. Cerebrovascular<br>Stroke (ischaemic), Stroke<br>3. Bone Disorders and<br>Fractures<br>Osteomyelitis, ONJ, | Joint Disorders<br>Osteoarthritis, Spinal<br>osteoarthritis, Arthritis,<br>Monoarthritis |
|  | extremity, Limb discomfort, Limb pain, Backache, Muscle spasms, Myalgia<br>3. Electrolyte and Metabolic Disorders<br>Hypocalcaemia, Hypercalcaemia, Hypophosphatemia, Dyslipidemia, Hyperlipemia, Elevated blood glucose<br>4. Liver Function–Related<br>Abnormal hepatic function, Abnormal liver function, Blood alkaline | 3. Oral and Disorders<br>Oropharyngeal pain, Dental caries, Tooth fracture, Periodontitis, Stomatitis<br>4. Gastrointestinal<br>Diarrhea, Nausea, Vomiting<br>5. Cardiovascular<br>Palpitation, Atrial fibrillation, Hypertension<br>6. Neurological and General<br>Headache, Dizziness, Malaise |  | Fracture, Fracture nonunion/delayed union, Fracture subtroch or femoral shaft, Hip fracture, Non-hip femur fracture, Femur fracture (all) |  |
|  | phosphatase is reduced<br>5. Skin Manifestations<br>(Allergy-Related)<br>Rash, Allergic dermatitis,<br>Urticaria<br>6. Neurological and<br>General Symptoms<br>Headache, Dizziness,<br>Peripheral oedema,<br>Hypertension<br>7. Laboratory<br>Abnormalities (Others)<br>Elevated serum creatine<br>phosphokinase, Keratitis | 7. Dermatologic and<br>Allergic<br>Eczema<br>8. Injection Site<br>Reactions<br>Injection site bruising,<br>Injection site erythema,<br>Injection site swelling,<br>Injection site pain,<br>Contusion<br>9. Laboratory<br>Abnormalities (Urine)<br>Blood urine present |  |  |  |

Eight studies^32–39^ evaluated the safety profiles of pharmacologic treatments for osteoporosis, including bisphosphonates, denosumab, teriparatide, abaloparatide, and romosozumab. Across randomized controlled trials and cohort studies, common adverse events included upper respiratory infections, musculoskeletal pain, dizziness, headache, and gastrointestinal disturbances. Injection site reactions and dermatologic symptoms (e.g., rash, urticaria) were reported particularly with subcutaneously administered agents. Laboratory abnormalities such as hypocalcemia, hypercalcemia, and liver enzyme alterations were noted, though typically transient and clinically manageable.

Serious adverse events were infrequent but observed across multiple classes. Long-term bisphosphonate use, for example, was associated in one study with an elevated risk of subtrochanteric or femoral shaft fractures after five or more years of use. Denosumab trials reported rare occurrences of hypocalcemia and potential risks of atypical infections. Cardiovascular events including atrial fibrillation and myocardial infarction were reported in trials of romosozumab and teriparatide, though causality remains uncertain.

Taken together, while the overall incidence of serious harms was low, the spectrum of mild to moderate adverse effects was broad and varied by drug class.

## Discussion

This systematic review provides an updated synthesis of evidence regarding osteoporosis screening and pharmacologic treatment, with specific relevance to the Japanese population. Overall, screening was generally associated with a modest reduction in hip and major osteoporotic fractures among older women. Risk assessment tools, including FRAX, demonstrated moderate predictive accuracy, though diagnostic accuracy varied depending on implementation strategy and population subgroup. Harms related to screening were infrequently reported. Notably, our review offers key updates to KQ2 and KQ5 by incorporating recent Japanese-language studies and population-specific considerations, while findings related to KQ4 reinforce previously established evidence.

Findings from KQ1 indicate that in the two included studies^12–13^, the effectiveness of osteoporosis screening was not solely dependent on the presence of screening protocols, but rather on the completion of the full cascade—including diagnostic follow-up and initiation of treatment^12^. Specifically, the impact of screening appeared to vary depending on whether DXA testing and subsequent clinical management were appropriately carried out following initial risk assessment^13^. Previous study^40^ further suggested that such implementation factors could influence outcomes. Additionally, the evidence questions the effectiveness of uniform, population-wide screening strategies, highlighting instead the need to consider individual characteristics and patient engagement behaviors. Several sources emphasized the importance of establishing supportive systems after screening and maintaining patient engagement, which may be critical considerations for future implementation efforts.

Among the studies^13–27^ included under KQ2a to KQ2c, strategies that combined clinical risk assessment tools with BMD testing were reported to be associated with improved diagnostic accuracy compared to the use of either modality alone in stratifying osteoporotic fracture risk. While FRAX was particularly effective when combined with BMD, several studies noted limitations in its discriminatory power among the very elderly, suggesting the need for predictive models that incorporate life expectancy, frailty, and competing mortality risks.

Some studies highlighted that the improved diagnostic performance of integrated approaches may be especially valuable in resource-limited settings for guiding treatment decisions, and may also aid in refining risk stratification among individuals at intermediate clinical risk. Regarding model calibration, there was evidence that incorporating comorbidities commonly observed in Japanese populations—such as diabetes or cardiovascular disease—into risk algorithms could enhance predictive performance. These findings reflect a broader literature trend emphasizing the importance of developing and validating population-specific risk models.

Given that misclassification—whether overestimation or underestimation—can directly impact treatment selection and unnecessary medication exposure, several studies underscored the need to address challenges related to model integration and calibration. These considerations should inform the design of future research and the implementation of fracture risk assessment strategies in clinical practice.

Based on the findings from studies included in Key Questions 4 and 5, pharmacological interventions for osteoporosis have been reported in multiple RCTs to be associated with a reduced risk of fractures. However, the literature emphasizes the need to evaluate these interventions in light of individual patient characteristics and potential adverse events. Several RCTs have shown that agents such as bisphosphonates and denosumab are associated with reductions in vertebral fracture incidence, while abaloparatide and romosozumab have demonstrated improvements in BMD with sequential use. These findings, drawn from trials such as the FRAME and ACTIVE-J studies, contribute to considerations regarding their applicability in Japanese populations.

Regarding adverse events, potential risks have been reported to vary depending on the type of drug and treatment duration. Although severe adverse events were generally rare, some studies reported atypical femoral fractures and osteonecrosis of the jaw associated with long-term bisphosphonate use, as well as cardiovascular events during romosozumab treatment. These findings highlight the importance of monitoring in high-risk populations and underscore the need for pre-treatment risk stratification. It should be noted that causal relationships in many of these reports remain inconclusive and require cautious interpretation.

In addition, mild adverse events such as musculoskeletal pain, dizziness, and injection site reactions were relatively common, particularly with non-oral formulations, potentially affecting treatment acceptability and long-term adherence. These observations emphasize the importance of individualized treatment planning that accounts not only for fracture risk and comorbidities but also for patient preferences and likelihood of treatment adherence over time.

In summary, the available evidence supports the clinical efficacy of pharmacological agents approved in Japan for osteoporosis treatment and underscores the necessity of shared decision-making that integrates both efficacy and safety profiles.

Several limitations should be noted. First, while the review incorporated Japanese-language studies, there remains a relative paucity of large-scale, Japan-specific RCTs on screening and treatment. This may limit generalizability, especially regarding real-world implementation in Japanese primary care. Second, most studies of risk assessment tools relied on AUCs without adequate reporting of calibration or reclassification metrics, which are critical for evaluating clinical utility. Finally, observational studies included in KQ5 varied in design quality, and residual confounding cannot be excluded.

In conclusion, this review identified clinical evidence supporting a two-step screening strategy— combining risk assessment tools with bone mineral density testing—particularly in older women at moderate to high risk. For pharmacological treatments approved in Japan, the reported frequency of serious adverse events was relatively low, and multiple studies have suggested their effectiveness in reducing fracture risk.

## Data Availability

All data supporting the findings of this systematic review are available upon reasonable request to the corresponding author.

## Disclaimer

This review was conducted independently of any funding organization. The findings and conclusions expressed herein do not represent the official positions of the authors’ affiliated institutions.

## Type of contribution of the authors

All authors were involved in the design and methodology of the review, data extraction and synthesis, and interpretation of the results. Each author contributed to the writing and critical revision of the manuscript, and all authors approved the final version and took responsibility for the integrity of the work.

## IRB Approval Code and Name of the Institution

Ethical review was deemed unnecessary by The Ethical Committee of Kurume University (health care & medical ethics), as this work is based solely on a literature review and does not involve human subjects or identifiable personal data.

## Acknowledgments

This review was conducted as part of the JPPSTF Project, which was commissioned by EVIDENCE STUDIO, a general incorporated association whose purposes include optimizing public healthcare expenditures in Japan. The contract was formally established with Kurume University, with which Kei Mukohara, the chair of the JPPSTF, is affiliated.

## Use of AI-assisted tools

During the preparation of this manuscript, an AI-assisted language model (ChatGPT, OpenAI, accessed in 2025) was used to assist in editing and improving the clarity and structure of the text. All content was critically reviewed and finalized by the authors, who take full responsibility for the accuracy and integrity of the manuscript.

## Search strategies for PubMed

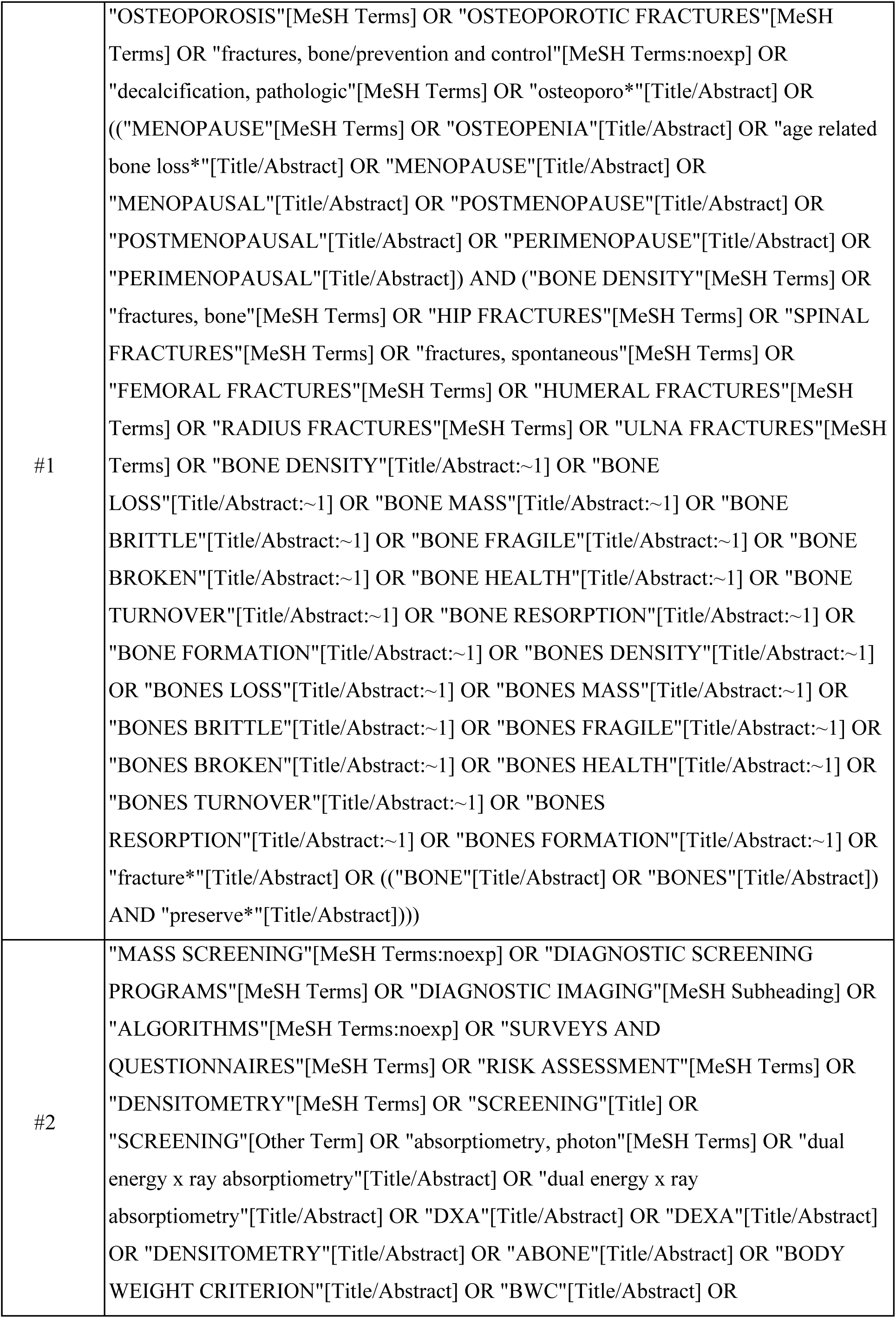
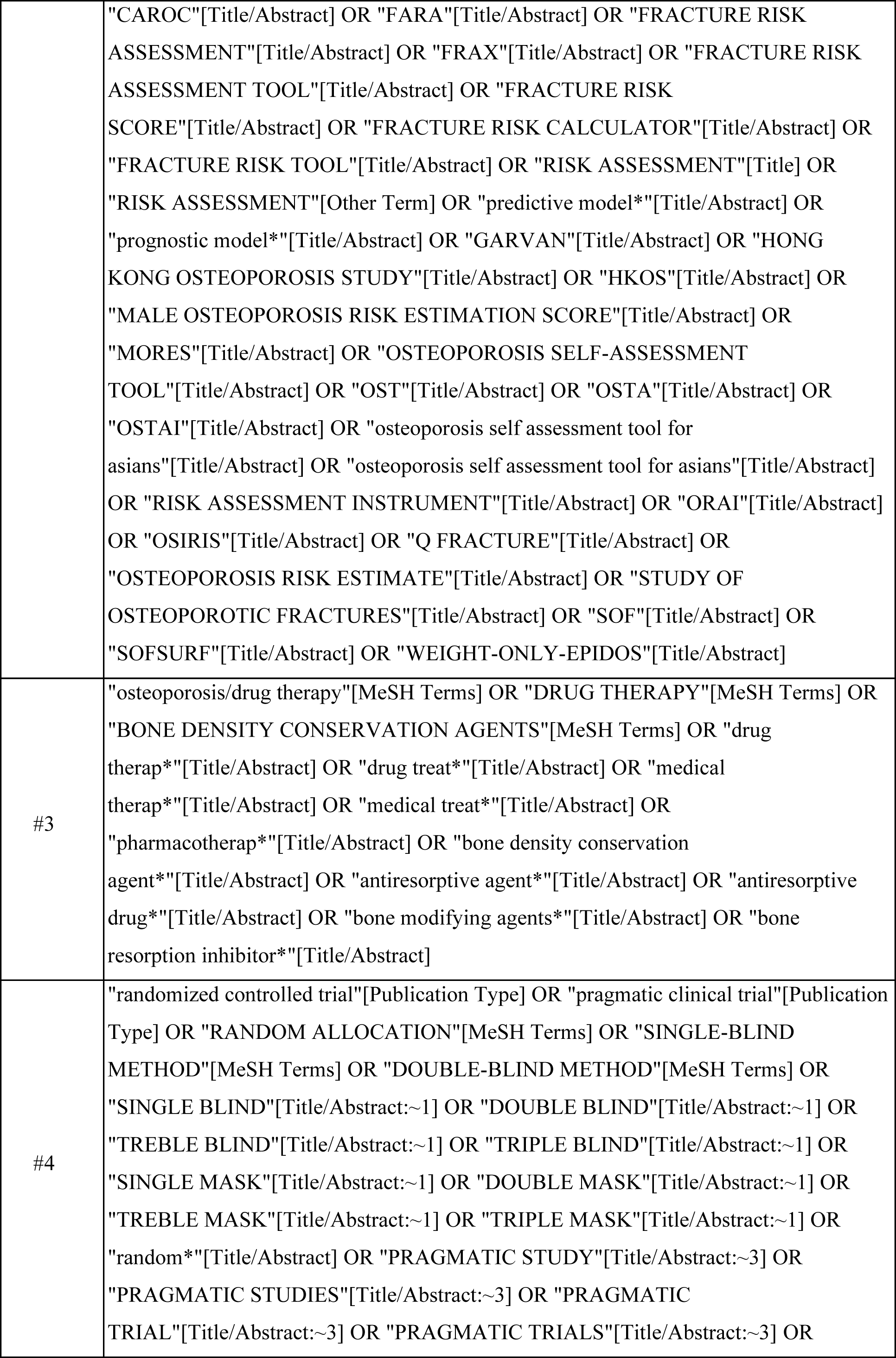
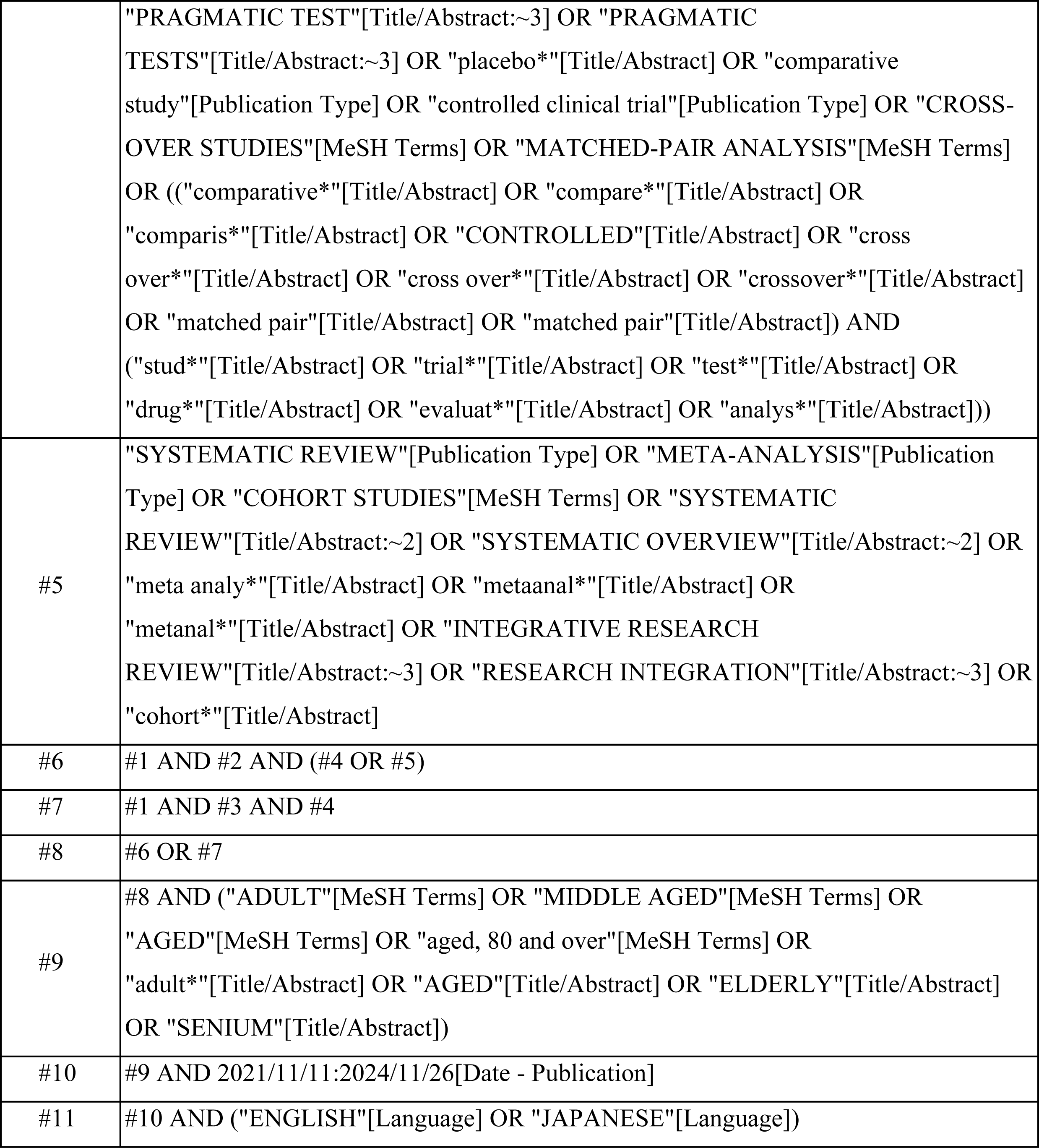

## Search strategies for Cochrane Library

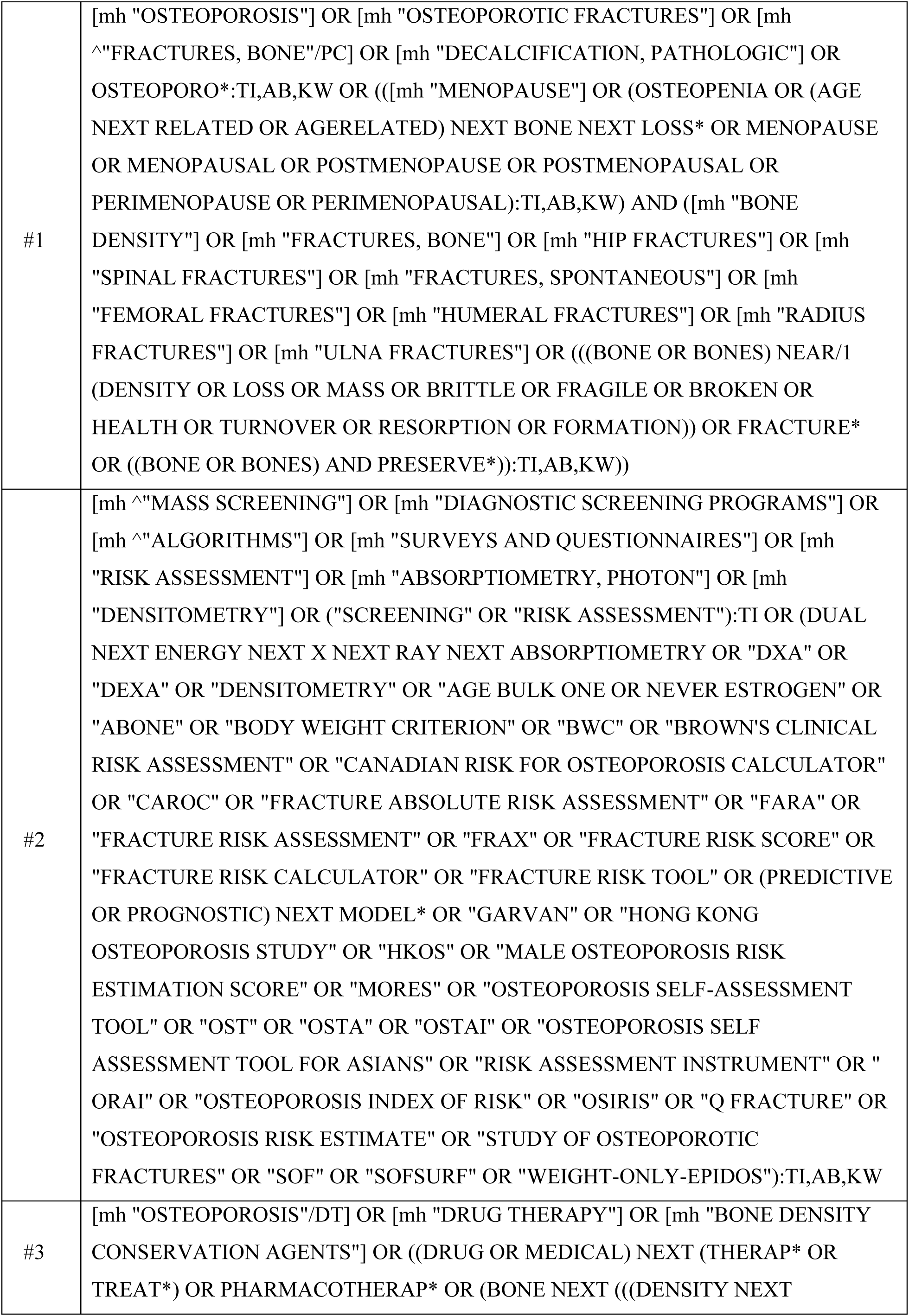
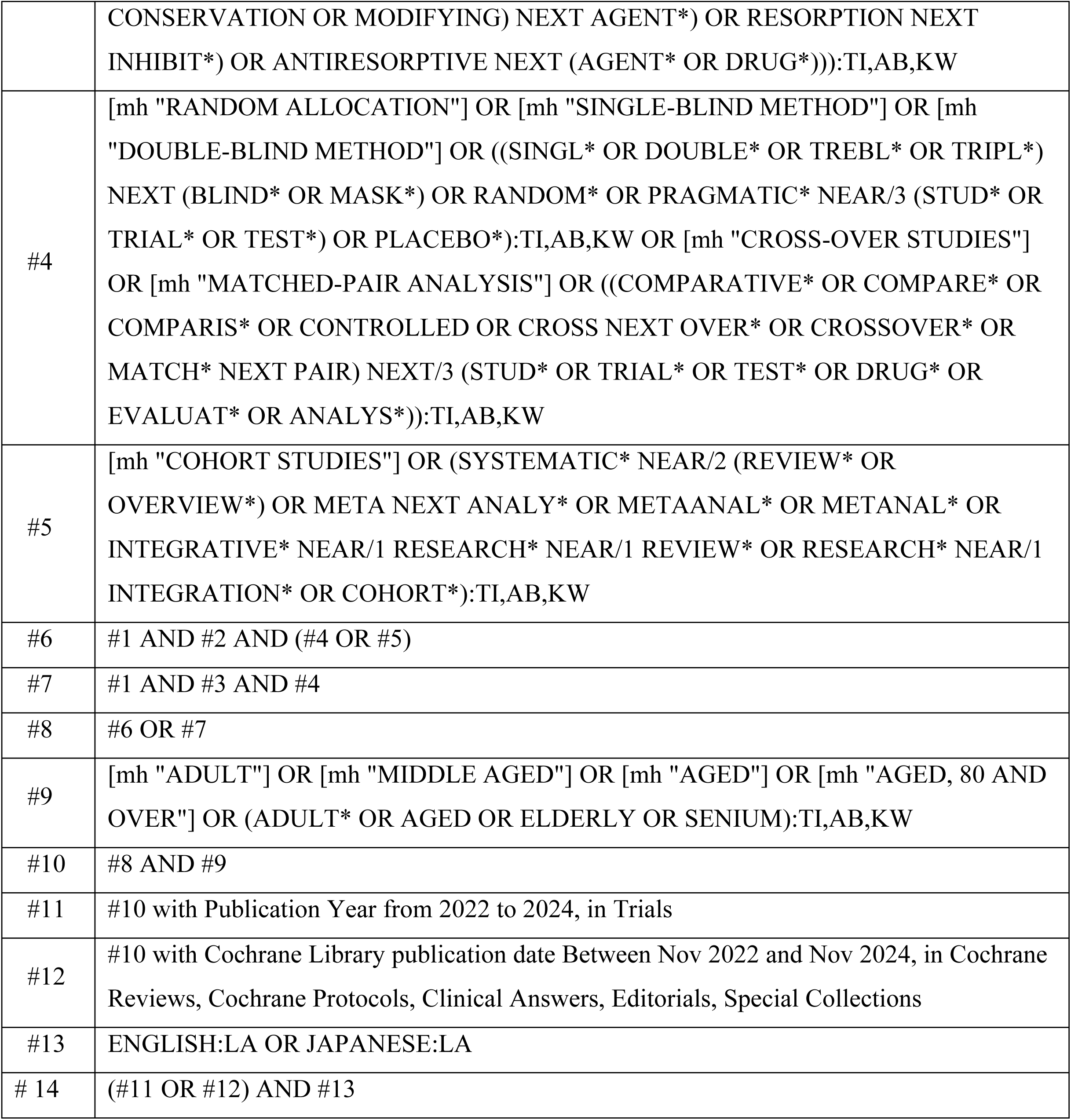

## Search strategies for Ichushi-Web (Japan Medical Abstracts Society)

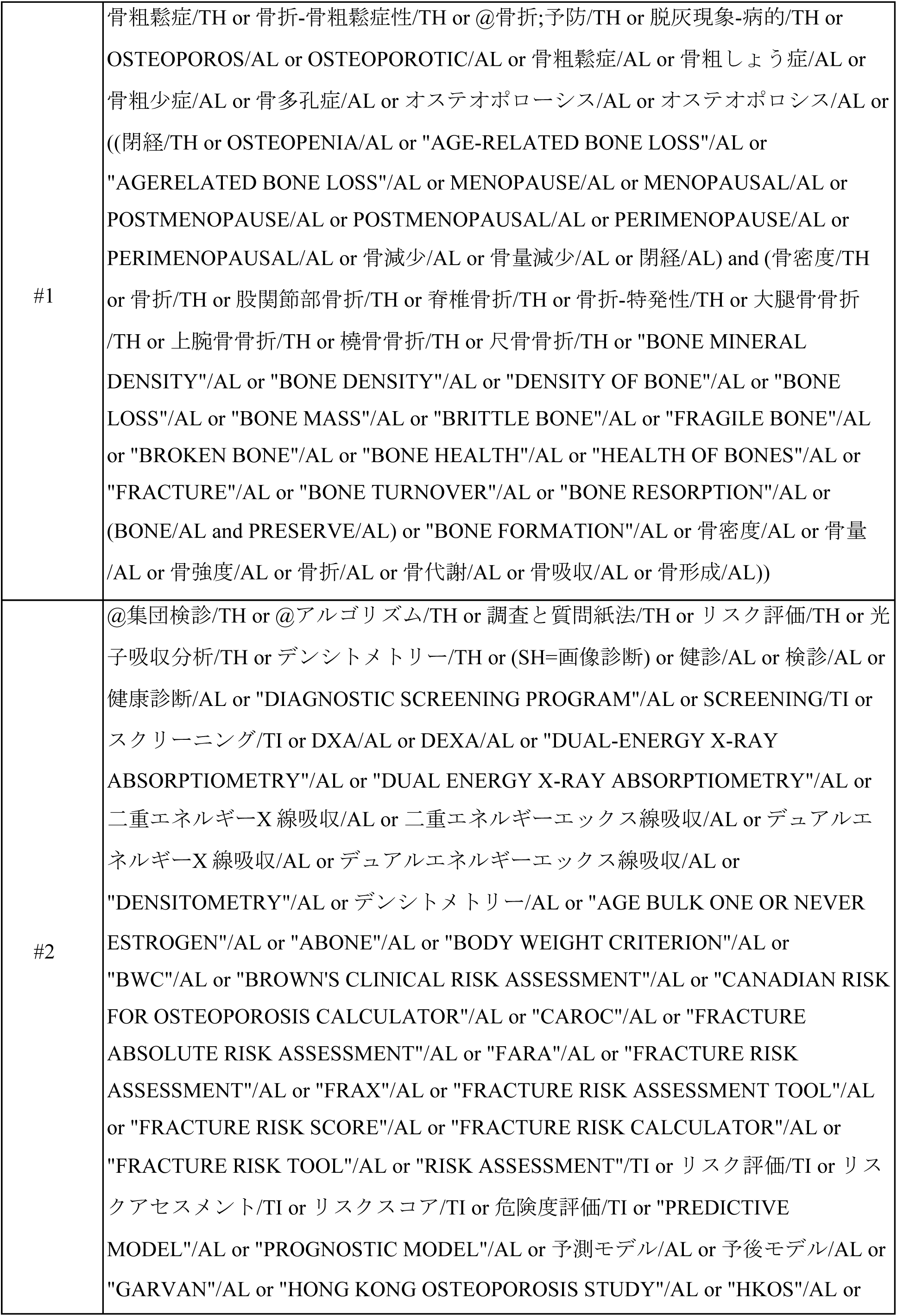
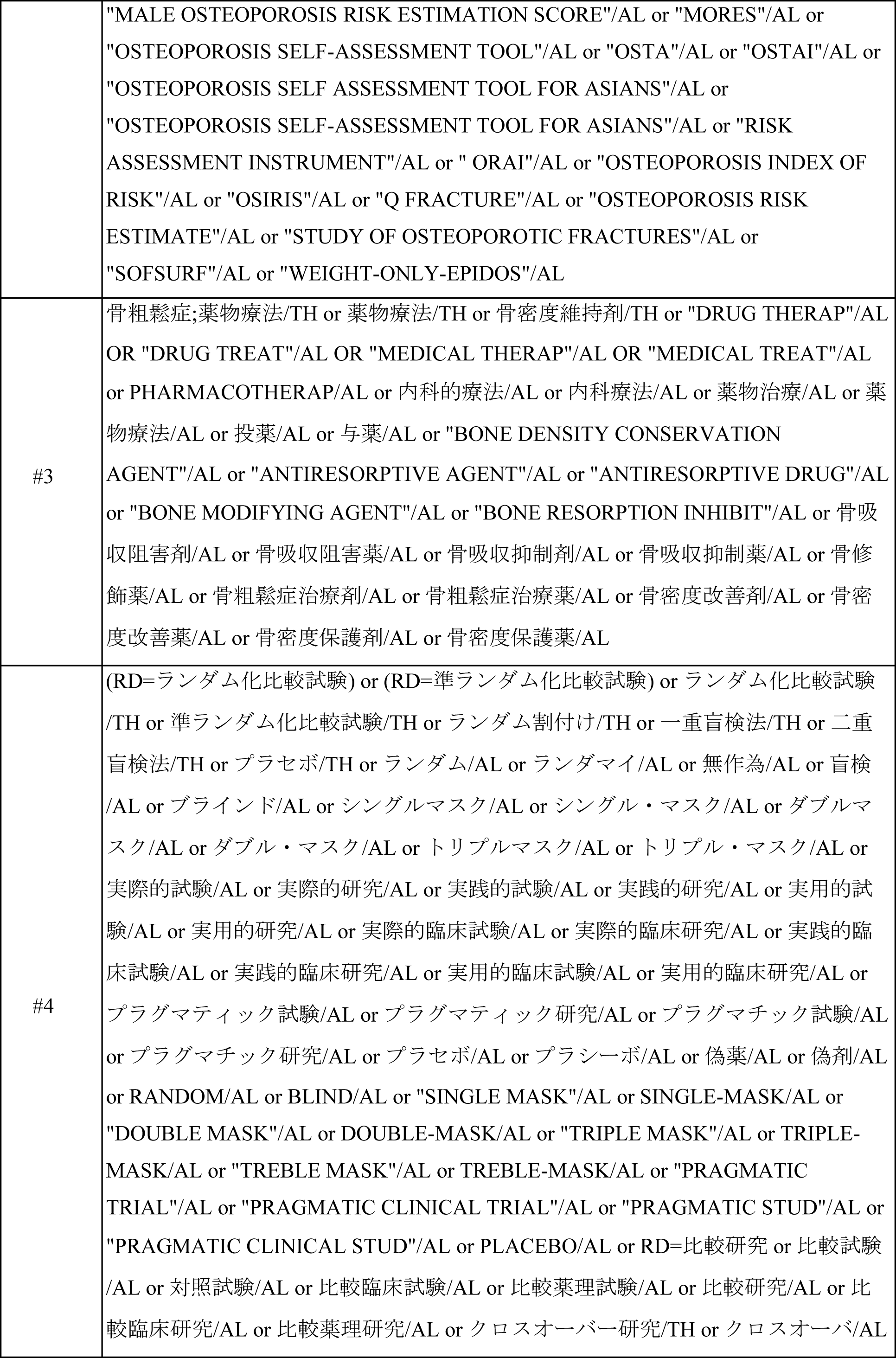
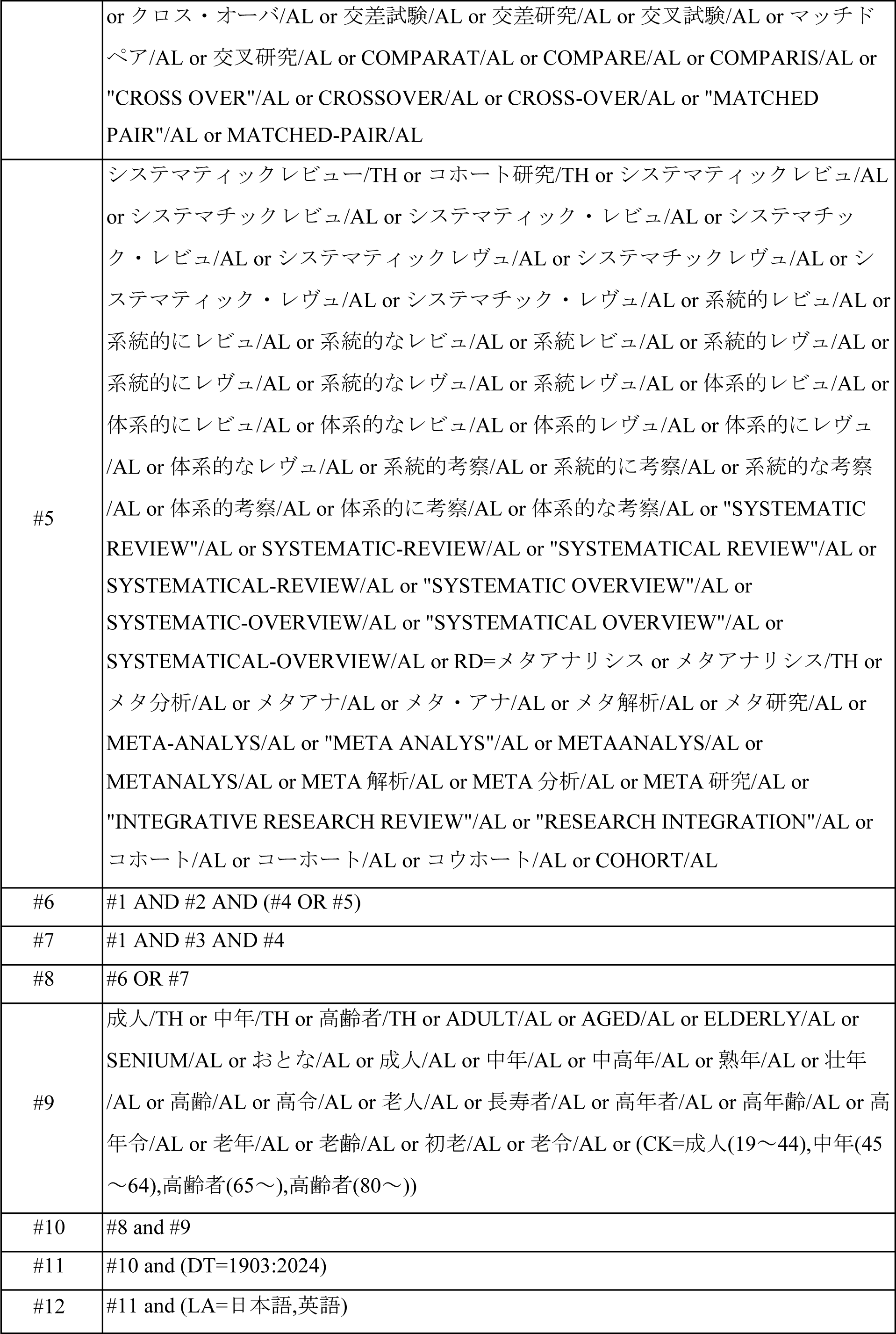

